# Concentration of SARS-CoV-2 from large volumes of raw wastewater is enhanced with the inuvai R180 system

**DOI:** 10.1101/2021.07.21.21260907

**Authors:** Silvia Monteiro, Daniela Rente, Mónica V. Cunha, Tiago A. Marques, Eugénia Cardoso, Pedro Álvaro, João Vilaça, Jorge Ribeiro, Marco Silva, Norberta Coelho, Nuno Brôco, Marta Carvalho, Ricardo Santos

## Abstract

Wastewater-based epidemiology (WBE) for severe acute respiratory syndrome Coronavirus 2 (SARS-CoV-2) is a powerful tool to complement syndromic surveillance: first, as an early-warning system for the spread of the virus in the community, second, to find hotspots of infection, and third, to aid in the early detection and follow-up of circulating virus variants.

Although detection of SARS-CoV-2 in raw wastewater may be prompted with good recoveries during periods of high community prevalence, in the early stages of population outbreaks concentration procedures are required to overcome low viral concentrations. Several methods have become available for the recovery of SARS-CoV-2 from raw wastewater, generally involving filtration. However, these methods are limited to small sample volumes, possibly missing the early stages of virus circulation, and restrained applicability across different water matrices. The aim of this study was thus to evaluate the performance of three methods enabling the concentration of SARS-CoV-2 from large volumes of wastewater: i) hollow fiber filtration using the inuvai R180, with an enhanced elution protocol and polyethylene glycol (PEG) precipitation; ii) PEG precipitation; and iii) skimmed milk flocculation. The performance of the three approaches was evaluated in wastewater from multiple wastewater treatment plants (WWTP) with distinct singularities, according to: i) effective volume; ii) percentage of recovery; iii) extraction efficiency; iv) inhibitory effect; and v) the limits of detection and quantification (The inuvai R180 system had the best performance, with detection of spiked controls across all samples, average recovery percentages of 64% for SARS-CoV-2 control and 68% for porcine epidemic diarrhea virus (PEDV), with low variability.

The inuvai R180 enables the scalability of volumes without negative impact on the costs, time for analysis, and recovery/inhibition. Moreover, hollow fiber filters favor the concentration of different microbial taxonomic groups. Such combined features make this technology attractive for usage in environmental waters monitoring.

## 1. Introduction

Surveillance of wastewater for epidemiological purposes has been previously used in public health, with the most important and successful example being the polio eradication program (GPEI, 2021). Given the ongoing Coronavirus disease 2019 (COVID-19) pandemic and accumulated reports of the presence of the severe acute respiratory syndrome coronavirus 2 (SARS-CoV-2) RNA in the stools of infected people and in raw wastewater (Gonzalez *et al*., 2020; Medema *et al*., 2020; Randazzo *et al*., 2020) the use of this matrix as a tool to monitor the emergence, prevalence, molecular epidemiology, and eventual phase out of SARS-CoV-2 in the community was prompted. Wastewater-based epidemiology (WBE) of SARS-CoV-2 has thus been gaining track among scientists, stakeholders, and decision makers throughout the world to complement syndromic surveillance and clinical testing. Although detection of SARS-CoV-2 may be performed directly on raw wastewaters with increased recovery percentages, ultimately optimization of concentration procedures is necessary in the early stages of virus circulation wherein low concentrations are expected (Gonzalez *et al*., 2020). Therefore, cost-effective, rapid and efficient concentration methods are required for monitoring SARS-CoV-2 or any other pathogen in raw wastewater for the successful deployment of WBE.

Existing methods for the recovery of viruses were primarily developed for the detection of nonenveloped viruses. Knowledge gaps concerning the recovery efficiencies of enveloped viruses, such as SARS-CoV-2, remain. A study by Haramoto *et al*. (2009) showed recovery efficiencies to be largely different for both types of viruses, with methods performing better for the recovery of nonenveloped viruses. Blanco *et al*. (2019) determined similar recovery efficiencies using precipitation with 20% polyethylene glycol (PEG) following glass wool concentration for enveloped (Transmissible gastroenteritis virus (TGEV)) and nonenveloped viruses (Hepatitis A virus (HAV)). A recent study by Ahmed *et al*. (2020) showed recovery efficiencies varying between 26.7 and 65.7% for murine hepatitis virus (MHV) in raw wastewater with very disparate recovery rates, even for similar methods, for this SARS-CoV-2 surrogate. Data using porcine epidemic diarrhea virus (PEDV) and aluminum flocculation-based concentration demonstrated recovery efficiencies of 11 and 3% for raw and treated wastewater, respectively (Randazzo *et al*., 2020).

Despite scarce information on diagnostic performance, SARS-CoV-2 RNA has been detected globally in raw wastewater with different approaches. Reported methods included ultrafiltration (Bertrand *et al*., 2021; Medema *et al*., 2020), ultracentrifugation (Wurtzer *et al*. 2020), PEG precipitation (Chavarria-Miró *et al*., 2020; La Rosa *et al*., 2020), aluminum flocculation (Randazzo *et al*., 2020), skimmed milk flocculation (Philo *et al*., 2021), and filtration through an electronegative membrane (Gonzalez *et al*., 2020; Haramoto *et al*., 2020).

In the present study, we evaluated the efficiency of SARS-CoV-2 recovery from raw wastewater using three concentration methods: i) a newly developed hollow-fiber filter, inuvai R180 (inuvai, a division of Fresenius Medical Care), with an improved elution protocol; ii) PEG precipitation; and iii) skimmed milk flocculation. The inuvai R180 filter has a large membrane area (1.8 m^2^) and a fiber inner diameter of 220 μm, allowing for the concentration of large volumes of water, including wastewater, without problems such as clogging or compromising of the membrane structure. The performance of the three methods was compared in aged raw wastewater according to several characteristics, including: i) effective volume tested; ii) frequency and consistency of detection; iii) percentage of recovery; iv) extraction efficiency; v) inhibitory effect on reverse transcription-qPCR (RT-qPCR); and vi) concentration information (including, Limit of Detection (LoD) and Limit of Quantification (LoQ)). This study benchmarks new and old methodologies for the detection of SARS-CoV-2 from raw wastewater for WBE applications.

## 2. Materials and Methods

### 2.1. SARS-CoV-2 control

SARS-CoV-2 control (nCoV-ALL-Control plasmid, Eurofins Genomics, Germany) was seeded into raw wastewater samples collected from five different WWTP in Portugal (as described below), following quantification by reverse transcription digital PCR (RT-dPCR) using two assays from the Charité protocol (Corman *et al*., 2020): E_Sarbecco and RdRp assays (Supplementary Table S1). Following absolute quantification (as described below), a stock solution with the concentration of 2.27 × 10^4^ genome copies per liter (GC/L) final concentration of wastewater (as measured for the E_Sarbecco assay) was prepared in DNase/RNase free water. The same stock was used for all experiments described below.

### 2.2. Porcine Epidemic Diarrhea Virus (PEDV) strain and cell lines

Porcine Epidemic Diarrhea Virus (PEDV) strain CV777 (kindly provided by Dr. Gloria Sanchez, IATA-CSIC) is an enveloped virus from the genus *Alphacoronavirus* and member of the *Coronaviridae* family, responsible for the porcine epidemic diarrhea. PEDV was propagated in Vero cell line (ATCC CCL-81, LGC Standards). Briefly, Vero cells were grown in Dulbecco’s Modified Eagle’s Medium (DMEM; Gibco), supplemented with 100 units/mL of penicillin (Lonza), 100 units/mL of streptomycin (Lonza), and 10% heat-inactivated fetal bovine serum (Biological Industries). Cells were cultured in T175 flasks at 37 (± 1) ºC under 5 % CO2. For infection with PEDV, cells were grown in T25 flasks and inoculated with 100 μL of viral stock. At 2h post infection, DMEM supplemented with 0.3% tryptose phosphate broth, 100 units/mL of penicillin (Lonza), 100 units/mL of streptomycin (Lonza), and 10 μg/μL trypsin, was added to the flasks. Flasks were then incubated at 37 (± 1) ºC in 5% CO2 for 4 days. PEDV were recovered following three cycles of freeze/thawing and centrifugation at 1,100 xg for 10 min. Quantification was performed by RT-dPCR as described on section 2.5 using the primers and probes from Supplementary Table S1 (Zhou *et al*., 2017), following nucleic acid extraction as described on section 2.4. After absolute quantification by RT-dPCR (as described below), a stock solution was prepared in DNase/RNase free water to obtain a PEDV final concentration of 1.21 × 10^4^ GC/L in wastewater. The same stock was used in all experiments described below.

### 2.3. Wastewater sample preparation

Twenty-four-hour composite samples were collected, on two separate rounds, from five wastewater treatment plants (WWTP) in Portugal (Serzedelo, Gaia, Alcântara, Beirolas and Guia). The first round comprised samples collected between April 27 and May 8, 2020 (*n* = 8; *n* = 2 for Serzedelo, Gaia and Beirolas; *n* = 1 for Alcântara and Guia) and the second round comprised samples collected between July 6-10, 2020 (*n* = 8; *n* = 2 for Serzedelo, Gaia and Guia; *n* = 1 for Alcântara and Beirolas). In each round, the samples were transported to the laboratory, refrigerated and within eight hours of collection. Samples collected in April-May were seeded with SARS-CoV-2 control whereas samples collected in July were seeded with PEDV. Raw wastewater samples were kept at 37 (± 1) ºC for seven days to ensure that the levels of SARS-CoV-2 RNA, if and where existing, decreased substantially prior to analysis. SARS-CoV-2 control and PEDV were seeded at concentrations of 2.27 × 10^4^ GC/L and 1.21 × 10^4^ GC/L, respectively (quantified as described previously).

Seeded raw wastewater samples were aliquoted and concentrated using three methods: (i) hollow fiber with the newly developed inuvai R180 filters (inuvai, a division of Fresenius Medical Care, Germany) followed by PEG precipitation (method 1); (ii) direct PEG precipitation (method 2); and (iii) skimmed-milk flocculation (method 3). All methods were tested using the same initial volume of wastewater (1-L) for a more accurate comparison.

Method 1 employed the use of hollow fiber filters: 1-L of raw wastewater was filtered through inuvai R180 filters using a peristaltic pump with a flow rate of 250 mL/min. The elution was performed in three steps: (i) air forward push using 60 mL of air; (ii) backflush with 250 mL of elution buffer (1× PBS with 0.01% NaPP and 0.01% Tween 80/0.001% antifoam) at a flow rate of 140-280 mL/min; and (iii) forward flush using 50 mL of elution buffer. The final elution volume was 300 mL. Samples were further concentrated by precipitation with 20% (w/v) PEG 8000 overnight (Blanco *et al*., 2019). Samples were centrifuged at 10,000 ×g for 30 min, the supernatant discarded, and the pellet resuspended in 5 mL 1× PBS, pH 7.4.

Method 2 used PEG precipitation: 20% PEG 8000 was added directly to 1-L of raw wastewater, with overnight precipitation followed by centrifugation as described above for method 1. Method 3 employed skimmed milk flocculation, performed in accordance with Calgua *et al*. (2008). Briefly, a pre-flocculated solution of 1% (w/v) skimmed milk pH 3.5 was prepared in artificial seawater. The solution of skimmed milk was then added to a final concentration of 0.01% (w/v) to 1-L of previously acidified raw wastewater (pH 3.5). Samples were stirred for 8h at room temperature and flocs were allowed to sediment for another 8h. Supernatant was carefully removed without disturbing the sediment. The final volume (approximately 500 mL) was centrifuged at 7,000 ×g for 30 min at 12 ºC. The supernatant was carefully discarded, and the pellet resuspended in 0.2 M phosphate buffer at pH 7.5 to a final volume of 5 mL. All concentrates were stored at – 80 (± 10) ºC until further analysis.

### 2.4. Nucleic acid extraction

Nucleic acid extraction was conducted using the QIAamp Fast DNA Stool mini kit (QIAGEN, Germany) from 220 μL of PEDV stock or concentrated raw wastewater samples according to the manufacturer’s instructions, recovering the nucleic acids in a final volume of 100 μL. Recovery efficiency for extraction was performed using Murine Norovirus 1 (MNV-1), added to the concentrates, as an extraction control. MNV was quantified using the assay described by Baert *et al*., 2008. Primers and probe information is provided on Supplementary Table S1. The extraction efficiency was calculated as

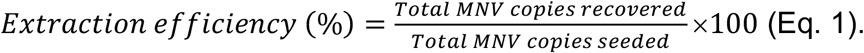

Following extraction, samples were stored at −30 (± 5) ºC until further processing.

### 2.5. Absolute quantification by RT-dPCR

RT-dPCR was used to determine the exact concentration of SARS-CoV-2 and PEDV spiked controls. Controls were amplified using the AgPath-ID One-Step RT-PCR kit (Thermo Fischer Scientific) with the set of primers and probe described on Supplementary Table S1 (PEDV; E_Sarbecco and RdRP assays). The 15 μL reaction mixture consisted of 7.5 μL of 2× RT-PCR buffer, 0.6 μL of 25× RT-PCR enzyme mix, 800 nM of each primer, 200 nM of probe, 3.63 μL RNase/DNase-free water, and 3 μL of DNA (diluted 4-, 5-, 6-fold). The reaction mixture was then spread over the QuantStudio 3D Digital PCR chip (Thermo Fischer Scientific) and the chips transferred to the QuantStudio 3D Digital PCR thermal cycler. Amplification was performed as follows: i) SARS-CoV-2: 10 min at 45 ºC, 10 min at 96 ºC, 39 cycles of 2 min at 58 ºC and 30 s at 98 ºC, and final elongation step for 2 min at 58 ºC; ii) PEDV: 10 min at 45 ºC, 10 min at 96 ºC, 39 cycles of 2 min at 60 ºC and 30 s at 98 ºC, and a final elongation step for 2 min at 60 ºC. Reactions were performed in duplicate, and a non-template control (NTC) was included in each run.

### 2.6. Relative quantification of seeded material in wastewater

Relative quantification of SARS-CoV-2 control, PEDV and MNV-1 was carried out by RT-qPCR on all extracts using the AgPath-ID One-Step RT-PCR kit (Thermo Fischer Scientific). The final volume of 25 μL was composed of 12.5 μL of 2× RT-PCR buffer, 1 μL of 25× RT-PCR enzyme mixture, 800 nM of each primer, 200 nM of the probe, 6.05 μL RNase/DNase-free water, and 5 μL of RNA. All RT-qPCR reactions were run on undiluted, 4- and 10-fold diluted extracts. RT-qPCR conditions were as follows: i) SARS-CoV-2 control: 10 min at 45 ºC, 10 min at 95 ºC, 45 cycles of 15 s at 95 ºC and 1 min at 58 ºC; ii) PEDV and MNV-1: 10 min at 45 ºC, 10 min at 95 ºC, 40 cycles of 15 s at 95 ºC and 1 min at 60 ºC. Standard curves, run with each PCR, for SARS-CoV-2 control (E_Sarbecco and RdRp assays), PEDV and MNV-1 were prepared in serial 10-fold dilutions in RNase/DNase-free water. Positive and NTC controls were also added to each PCR assay. Limits of detection (LoD) and quantification (LoQ) were determined in RNase/DNase-free water. The LoD was considered the lowest concentration of target that could be consistently detected (in more than 95% replicates tested) (Burd *et al*., 2010) and LoQ, the lowest concentration at which the performance of the method is acceptable, with a coefficient of variation below 35% (Klymus *et al*., 2020).

### 2.7. Recovery efficiency

The mean recovery efficiency of SARS-CoV-2 control and PEDV for each method was calculated using the copies quantified by RT-qPCR as follows (Eq. 2):

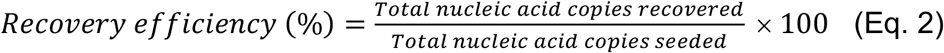

The mean and standard deviation for each method were also calculated.

### 2.8. Quality control

To minimize nucleic acid carry-over and cross-contamination, sampling concentration, extraction procedures and RT-qPCR/RT-dPCR were performed in separate rooms of the laboratory. A process blank and extraction blank were included for each concentration method and each nucleic acid extraction, respectively. As described above, and before spiking, all wastewater samples were aged to decay potentially present SARS-CoV-2 RNA; following aging, all spiked samples were tested in parallel with the corresponding unseeded samples to rule out or estimate the contribution of potentially native SARS-CoV-2 and PEDV.

### 2.9. Data analyses

All data analyses were performed with SPSS Statistics 26 (IBM). Repeated measurement ANOVA was conducted to compare the differences between the parameters estimated for the three methods. In all cases, p-values < 0.05 were considered statistically significant.

## 3. Results and discussion

### 3.1. Quantification of controls

Appropriate quantification of the controls used in spiking experiments and in standard curve for RT-qPCR is extremely important, as it will influence downstream data interpretation. That is why we opted for RT-dPCR, with high precision and sensitivity, for the absolute quantification of controls. Digital PCR works by partitioning a unique sample into thousands of individual reactions running in parallel, being particularly useful for low-abundance targets or targets in complex matrices. Through Poisson statistics, the total number of target molecules is calculated, with no need for external reference standards (Monteiro and Santos, 2017). Several dilutions of SARS-CoV-2 control and PEDV, in duplicate, were quantified by RT-dPCR. The concentrations of the initial stocks for SARS-CoV-2 control were 1.94 × 10^8^ GC/μL and 1.00 × 10^8^ GC/μL for E_Sarbecco and RdRp assays, respectively. Concentration of PEDV as determined by RT-dPCR was 1.20 × 10^8^ GC/μL.

### 3.2. Method comparison using SARS-CoV-2 and PEDV as surrogates for SARS-CoV-2

All unseeded wastewater samples were negative for the presence of SARS-CoV-2 and PEDV. Samples were chosen in periods with low number of daily COVID-19 cases (mean for entire country, 287 from April 27 to May 8, and 374, between July 6 and 10, 2020) (DGS, 2020). All process and extraction blanks were negative.

The effective volume tested within each method was the same (2.2 mL): all methods started with the same initial volume (1-L) of wastewater, followed by concentration steps prior to extraction and sediment resuspension in 5 mL of elution buffer; samples tested across the three methods were extracted using the same extraction protocol, and the same volumes and dilutions were analyzed by RT-qPCR. Nonetheless, the inuvai R180 filters (method 1) enabled the filtration of 2.5 – 5-L of raw wastewater. Increasing the initial volume of sample with the inuvai R180 filters would conduct to an increment of the effective volume assayed from 2.2 mL to 5.5 – 11 mL without further increases in the concentration time, the concentrate volume, costs for analysis, and RT-qPCR inhibition. On the other hand, increasing the volume of filtration in the skimmed milk flocculation method (and therefore, theoretically, increasing the effective volume assayed; method 3) would imply an increase of skimmed milk and artificial seawater, as well as of HCl to adjust the pH; the volume of concentrated matter and, therefore, of the concentrate would also increase, leading to a decrease in the efficiency of extraction and an increase of inhibitory effects on RT-qPCR. Additionally, increasing the processing volume would require the acquisition of larger volume sample containers, which would also take up more space in the laboratory. Concomitantly, increasing the processing volume when using solely PEG precipitation (method 2) implicates increasing substantially the volume to be centrifuged, which increases the time spent in the concentration step and the costs due to the usage of larger amounts of PEG.

SARS-CoV-2 control and PEDV were used to compare concentration recoveries. The highest average percentage of recovery was obtained with the inuvai R180 system at 64% (± 6%) for SARS-CoV-2 control and 68% (± 7%) for PEDV, with global recoveries varying between 50 and 82% (Fig. 1A).

**Fig. 1.**
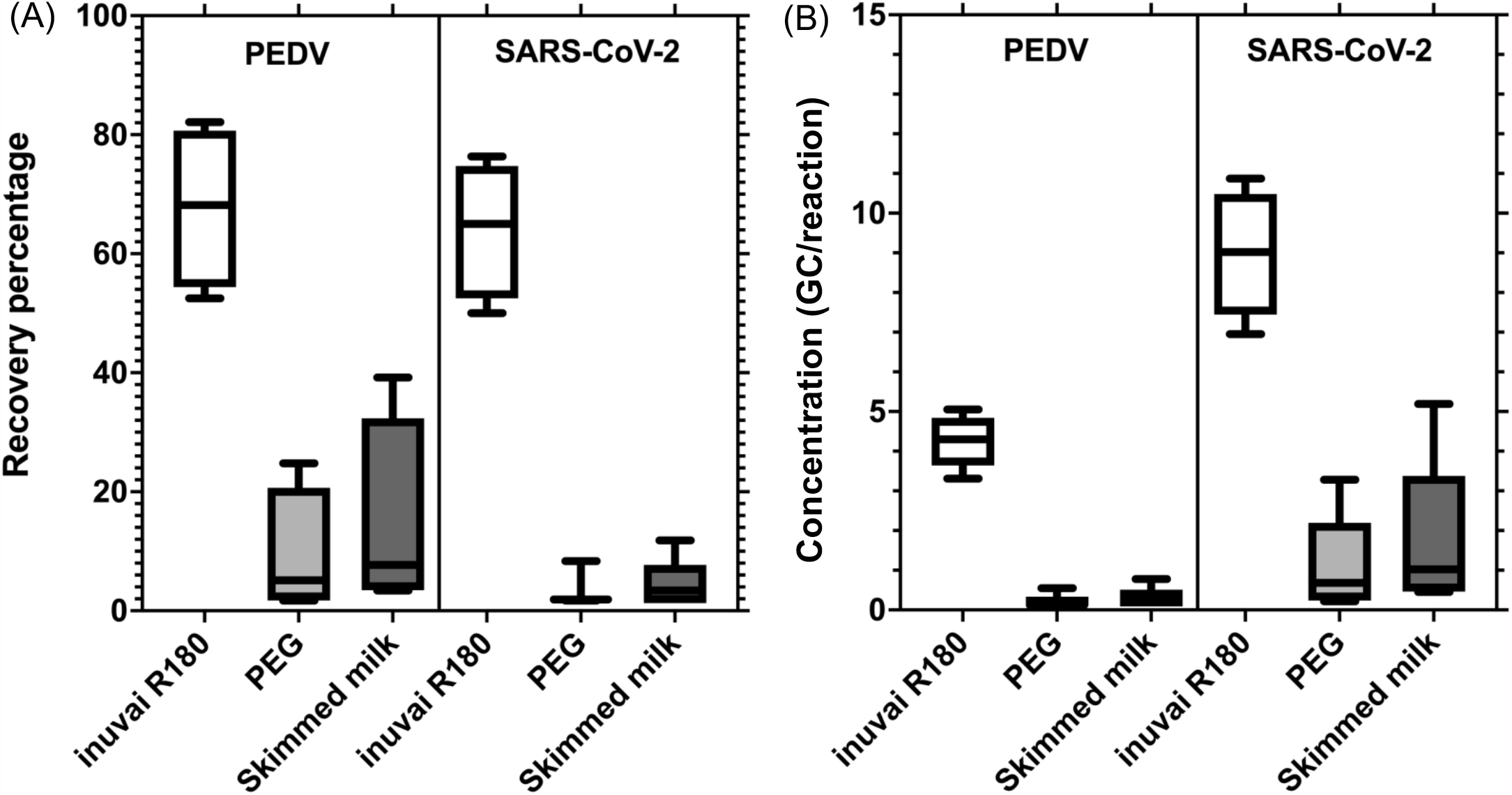
Performance of concentration methods for the detection of PEDV and SARS-CoV-2 control from raw wastewater. Percentage of recovery obtained in each method (A). log transformed concentration of viral genome copies detected by RT-qPCR in each method (B). The inuvai R180 system presented the highest average percentage of recovery and concentration, followed by PEG precipitation and skimmed milk flocculation.

PEG precipitation had the lowest percentage of recovery for PEDV (9% (± 5%)). Recovery with skimmed milk performed only slightly better (14% (± 8%)) (Fig. 1A). Recovery using SARS-CoV-2 control was similar for PEG and skimmed milk (4% (± 2%)). There were statistically significant differences in the lower recovery percentage of PEG and skimmed milk compared to inuvai R180 *(F*(1, 3) = 14.94, *p* = 0.03 for PEDV and *F*(1, 3) = 171.7, *p* = 0.006 for SARS-CoV-2 control).

The inuvai R180 was the single method that consistently led to nucleic acid detection in all samples. Concentration using PEG and skimmed milk led to the detection of PEDV in 50% of the samples, while detection of SARS-CoV-2 control was attained in 38% and 63% of the samples, respectively.

The method using the inuvai R180 system led to detection by RT-qPCR of the highest mean concentration of genome copies, for both targets: 8.98 and 4.25 GC/reaction for SARS-CoV-2 and PEDV, respectively. Concentration with PEG (1.19 and 0.21 GC/reaction for SARS-CoV-2 and PEDV, respectively) and skimmed milk (1.74 GC/reaction for SARS-CoV-2 and 0.28 GC/reaction for PEDV) showed similar results (Fig. 1B).

Our recovery values using the inuvai R180 system were similar to those reported for MHV, while enabling an increase in the filtration volume (Ahmed *et al*., 2020). For PEG precipitation and skimmed milk flocculation the recoveries were slightly higher than those reported by Philo *et al*. (2021). The authors used a concentration of 14% (w/v) of PEG compared to 20% (w/v) PEG in our study. The use of higher concentrations of PEG, although implying increased costs, has been shown to increase the recovery of enveloped viruses from 31% to 51% (Blanco *et al*., 2019). In our study, recovery values for PEG precipitation were higher than those reported by Pérez-Cataluña *et al*. (2021) when using similar nucleic acid extraction method (spin column). McMinn *et al*. (2021) developed a method for the recovery of coronavirus from raw wastewater also using hollow fibers as a primary concentration approach, followed by Concentrating Pipette Select™ (CP Select™), reporting overall recovery values for human coronavirus OC43 of 22%. Differences in recovery between our study and that of McMinn *et al*. (2021) may be attributed to the filter that used in our study (inuvai R180 vs Rexeed), coupled with an enhanced elution strategy with three steps that we adopted, and/or to the secondary concentration protocol. The inuvai R180 filter has a reduced nominal pore size (≤ 5.5 nm with a correspondent cut-off ≤ 18.8 Kda) compared to the Rexeed 15S, which has a more open pore structure. Additionally, the filter used in our study has a larger membrane area (1.8 m^2^ for inuvai R180 vs 1.5 m^2^ for Rexeed S15) and larger fiber inner diameters (220 μm for inuvai R180 vs 185 μm for Rexeed S15). In addition to the optimized elution and secondary concentration protocols, such features might help justify the differences registered in the recovery efficiencies of our study and McMinn *et al*. (2021).

The extraction efficiency using MNV as proxy averaged 70% (±19%) for inuvai R180 protocol. Extraction efficiencies for PEG precipitation and skimmed milk flocculation averaged 50% (±15%) and 36 (±13%), respectively.

Detection of SARS-CoV-2 control and PEDV using the inuvai R180 system was consistently achieved with the 1/4-fold dilution, while for undiluted spiked samples, only 38% could be detected without inhibition. PEG precipitation was the single method that detected both targets from undiluted samples, although inhibition still occurred (as evidenced subsequently by testing the 4- and 10-fold dilution). As for the skimmed milk concentration method, detection in undiluted concentrates was found for 75% of the samples, although inhibition still occurred (as measured by the dilutions). These results indicate that inhibitory effects exerted upon RT-qPCR could be confirmed for the three methods under comparison.

Overall, our results showed that the inuvai R180 system coupled with an improved elution protocol is highly suitable for the detection of SARS-CoV-2 and PEDV, exhibiting the highest percentage of detection and mean recovery value. Additionally, this method also showed greater extraction efficiency and larger volume processing without increased cost or time for downstream analyses. Furthermore, the performance of the inuvai system showed consistency across raw wastewater samples from different catchments / WWTP, including the Serzedelo WWTP, which is highly impacted by industrial effluents (tannery industry) and therefore an extremely complicated matrix to work with altogether, a result corroborated by the Pan-European Umbrella study (Gawik *et al*., 2021). In the Umbrella study, raw wastewater samples from different European countries were collected and sent for analysis in a centralized laboratory. In parallel, the same samples were also analyzed in each country for comparison of results. The centralized European laboratory was unable to recover SARS-CoV-2 RNA from Serzedelo raw wastewater presenting low recovery percentages (0.1%) and lower concentrations of crAssphage compared to the other samples analyzed. The same sample, analyzed by our group and using the inuvai R180 system, was positive for SARS-CoV-2 and the concentration of crAssphage was 3-log above that detected by the centralized laboratory. These results demonstrate the difficulty of working with this raw wastewater, highlighting the need to test method performance in raw wastewater from different origins.

### 3.3. RT-qPCR efficiency

After establishing the inuvai R180 system as gold-standard for primary concentration, the efficiency of the relative quantification method (RT-qPCR) was assessed by calculating the LoD and LoQ for the E_Sarbecco and RdRp assays using SARS-CoV-2 control. Fig. 2 displays the subset of points from the standard curve to determine the LoD and LoQ.

**Fig. 2.**
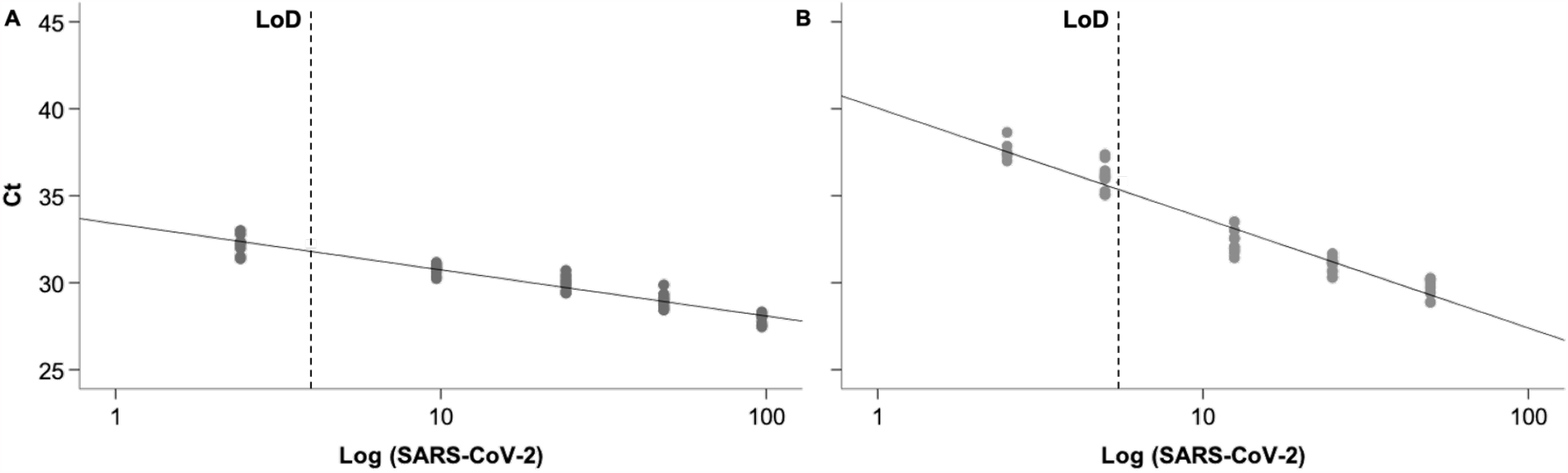
LoD for SARS-CoV-2 relative quantification assays. Subset of standard curve points used to determine the smallest concentration of SARS-CoV-2 detected by E_Sarbecco assay at a 95% confidence level (A). Curve to determine the smallest concentration of SARS-CoV-2 detected by RdRp assay at a 95% confidence level (B).

The LoD was 3.99 GC and 5.52 GC per reaction for the E_Sarbecco and RdRp assays, respectively. This corresponded to a method LoD of 2.73 × 10^3^ GC/L for E_Sarbecco and 3.79 × 10^3^ GC/L for RdRp using the inuvai R180 system.

As for the LoQ, the results were 66 GC and 178 GC per reaction for the E_Sarbecco and RdRp assays, respectively. This corresponded to a method LoQ of 4.56 × 10^4^ GC/L for E_Sarbecco and 1.22 × 10^5^ GC/L for RdRp assay.

The LoD obtained in our study were inferior to those obtained by Philo *et al*. (2021). Pérez-Cataluña *et al*. (2021) reported similar LoD for E_Sarbecco assay, while also presenting method-dependence LoD. Gonzalez *et al*. (2020), testing the CDC assay (N1, N2, and N3), reported different theoretical limits of detection depending on the RT-qPCR assay used but the LoD were similar to those obtained in our study. A comparison between the performance of our method (evaluated through LoD and LoQ) and the method reported by McMinn *et al*. (2021) would have been useful, given that the authors have also used hollow-fiber filters for primary concentration, but such parameter information is missing on the former report. In fact, information on LoQ is missing from most publications with very few exceptions, such as LaTurner *et al*. (2021) who, while testing five distinct concentration methods, reported LoQ ranging from 2.76 × 10^5^ to 8.39 × 10^6^ GC/L. Philo *et al*. (2021) calculated their LoQ in nuclease-free water to be 100 gene copies per reaction for all CDC assays.

### 4. Conclusions

Data from our study demonstrates the importance of validating concentration procedures using seeded controls. Although other studies have tested the efficiency of concentration and extraction methods, this study showed the stability of the inuvai R180 system for the recovery of seeded controls in raw wastewater from WWTP with different composition particularities, including effluents from the tannery industry. A single concentration method may not necessarily be ideal to be used in waters from different backgrounds. In this study, the inuvai R180 system with improved three-step elution protocol was selected for monitoring SARS-CoV-2 in raw wastewaters. Such system is attractive as it enables the concentration of large volumes of raw wastewater, while also being useful to concentrate larger volumes of samples from other origins, such as treated wastewater, environmental waters and drinking water. This feature enables handling a single concentration method across different water types without sensitivity loss, increasing costs or time for analysis, while also allowing a less challenging result comparison.

For an effective environmental surveillance to be put in place, not only for SARS-CoV-2 but also for potential future pandemics involving enveloped virus, it is paramount to have validated methods. Nonetheless, comparisons between published methods are difficult as they differ in many aspects including: i) seeding controls; ii) concentration methods; iii) extraction methods; iv) diagnostic and quantification molecular assays and genome targets; v) and mostly, the accepted performance levels. Some publications only mention the recovery efficiency (Ahmed *et al*., 2020; McMinn *et al*., 2021), others mention the recovery efficiency and the LoD but not LoQ (Gonzalez *et al*., 2020; Randazzo *et al*., 2020; Pérez-Cataluña *et al*., 2021), some mention LoQ but not LOD (LaTurner *et al*., 2021), while other studies show all data performance, including LoD, LoQ and recovery percentages (Philo *et al*., 2021). Additionally, different studies calculate the LoD and LoQ differently. The information collected from different studies should inform laboratories on method performance. A ‘one size fits all’ approach, that is having a single standardized method worldwide for the concentration of SARS-CoV-2, may not be the best approach. This was demonstrated with the Umbrella study (Gawik *et al*., 2021), due to several issues, including: (i) laboratories already have their own preferred methods with performances studied; (ii) the methods may not be useful for application in less economically developed countries; (iii) or simply because it is difficult to get a hold of laboratory materials/equipment (as it was the case of ultrafiltration filters or ultracentrifuges). Nonetheless, standards as to what should be asked in terms of method performance should be established so that laboratories could gather all the information about the methods to make a more informed choice. Wastewater surveillance has the potential to prevent the occurrence of new outbreaks (Peiser, 2020), and to help understand changes in the pandemic trends. Effective methods, with performance specifications detailed, are paramount for wastewater surveillance to be applied in accurately describing the transmission of SARS-CoV-2 in the community. This study expands the knowledge on analytical methods introducing a method with robust performance for SARS-CoV-2 detection in wastewater and establishing a step forward for the global application of WBE not only for this pandemic but also in future health crisis as the established protocol is modular for different taxonomic groups.

## Data Availability

All data contained in this manuscript has not been peer reviewed, published or accepted in any other platform

## CrediT authorship contribution statement

**Sílvia Monteiro**: conceptualization, methodology, software, validation, formal analysis, investigation, writing – original draft, writing – review and editing, visualization. **Daniela Rente**: investigation. **Mónica V. Cunha**: review and editing. **Tiago A. Marques**: review and editing. **Eugénia Cardoso**: review and editing, sampling; **Pedro Álvaro**: review and editing, sampling; **João Vilaça**: review and editing, sampling; **Jorge Ribeiro**: review and editing, sampling; **Nuno Brôco**: project administration, funding acquisition, review and editing; **Marta Carvalho**: project administration, funding acquisition, review and editing; **Ricardo Santos**: conceptualization, methodology, resources, formal analysis, writing – review and editing.

## Acknowledgements

We thank all the workers from Águas de Portugal Group who contributed with wastewater samples and those who contributed with critical discussion.

This work was funded by Programa Operacional de Competitividade e Internacionalização (POCI) (FEDER component) and Programa Operacional Regional de Lisboa (Project COVIDETECT, LISBOA-01-02B7-FEDER-048467).

Strategic funding of Fundação para a Ciência e a Tecnologia (FCT), Portugal, to cE3c, BioISI and CEAUL Research Units (UIDB/00329/2020, UIDB/04046/2020 and UIDB/00006/2020) is gratefully acknowledged.

