## Supplementary_Table_1 for "Concentration of SARS-CoV-2 from large volumes of raw wastewater is enhanced with the inuvai R180 system"

Primers and probes used in this study

| Assay | Sequence (5' - 3')[a] | Length (bp) | Location in SARS-CoV-2 genome (bp) |
|---|---|---|---|
| MNV | F: CACGCCACCGATCTGTTCTG<br>R: GCGCTGCGCCATCACTC<br>P: 6FAM-CGCTTTGGAACAATG-MGB | 108 | 4,972 – 5,080 |
| PEDV | F: CAGGACACATTCTTGGTGGTCTT<br>R: CAAGCAATGTACCACTAAGGAGTGTT<br>P: FAM-ACGCGCTTCTCACTAC-MGB | 140 | 26,010 - 26,149 |
| SARS-CoV-2: E_Sarbecco | F: ACAGGTACGTTAATAGTTAATAGCGT<br>R: ATATTGCAGCAGTACGCACACA<br>P: 6FAM-ACACTAGCCATCCTTACTGCGCTTCG-BHQ | 112 | 26,141 – 26,253 |
| SARS-CoV-2: RdRp | F: GTGARATGGTCATGTGTGGCGG<br>R: CARATGTTAAASACACTATTAGCATA<br>P1: 6FAM-CCAGGTGGWACRTCATCMGGTGATGC-BHQ<br>P2: 6FAM-CAGGTGGAACCTCATCAGGAGATGC-BHQ | 99 | 15,361 – 15,460 |
| SARS-CoV-2: N_Sarbecco | F: CACATTGGCACCCGCAATC<br>R: GAGGAACGAGAAGAGGCTTG<br>P: 6FAM-ACTTCCTCAAGGAACAACATTGCCA-BHQ | 127 | 28,555 – 28,682 |

[a] W is A/T; R is G/A; M is A/C; S is G/C. FAM: 6-carboxyfluorescein; MGB: minor groove binder; BHQ: blackhole quencher.
